# Breast cancer risk based on a deep learning predictor of senescent cells in normal tissue

**DOI:** 10.1101/2023.05.22.23290327

**Authors:** Indra Heckenbach, Mark Powell, Sophia Fuller, Jill Henry, Sam Rysdyk, Jenny Cui, Amanuel Abraha Teklu, Eric Verdin, Christopher Benz, Morten Scheibye-Knudsen

**Affiliations:** Center for Healthy Aging, Department of Cellular and Molecular Medicine, University of Copenhagen, Copenhagen, Denmark; Buck Institute for Research on Aging, Novato, CA, USA; Zero Breast Cancer, San Rafael, CA, USA; Graduate Group in Biostatistics, School of Public Health, University of California, Berkeley, CA, USA; Susan G. Komen Tissue Bank at the IU Simon Comprehensive Cancer Center, Indiana University School of Medicine, Indianapolis, IN, 46202, USA; Department of Biochemistry and Molecular Biology, College of Health Sciences, Mekelle University, Ethiopia

## Abstract

**Background:** The ability to predict future risk of cancer development in non-malignant biopsies is poor. Cellular senescence has been associated with cancer as either a barrier mechanism restricting autonomous cell proliferation or a tumor-promoting microenvironmental mechanism that secretes pro-inflammatory paracrine factors. With most work done in non-human models and the heterogenous nature of senescence the precise role of senescent cells in the development of cancer in humans is not well understood. Further, more than one million non-malignant breast biopsies are taken every year that could be a major source of risk-stratification for women.

**Methods:** We applied single cell deep learning senescence predictors based on nuclear morphology to histological images of 4,411 H&E-stained breast biopsies from healthy female donors. Senescence was predicted in the epithelial, stromal, and adipocyte compartments using predictor models trained on cells induced to senescence by ionizing radiation (IR), replicative exhaustion (RS), or antimycin A, Atv/R and doxorubicin (AAD) exposures. To benchmark our senescence-based prediction results we generated 5-year Gail scores, the current clinical gold standard for breast cancer risk prediction.

**Findings:** We found significant differences in adipocyte-specific IR and AAD senescence prediction for the 86 out of 4,411 healthy women who developed breast cancer an average 4.8 years after study entry. Risk models demonstrated that individuals in the upper median of scores for the adipocyte IR model had a higher risk (OR=1.71 [1.10-2.68], p=0.019), while the adipocyte AAD model revealed a reduced risk (OR=0.57 [0.36-0.88], p=0.013). Individuals with both adipocyte risk factors had an OR of 3.32 ([1.68-7.03], p<0.001). Alone, 5-year Gail scores yielded an OR of 2.70 ([1.22-6.54], p=0.019). When combining Gail scores with our adipocyte AAD risk model, we found that individuals with both of these risk predictors had an OR of 4.70 ([2.29-10.90], p<0.001).

**Interpretation:** Assessment of senescence with deep learning allows considerable prediction of future cancer risk from non-malignant breast biopsies, something that was previously impossible to do. Furthermore, our results suggest an important role for microscope image-based deep learning models in predicting future cancer development. Such models could be incorporated into current breast cancer risk assessment and screening protocols.

**Funding:** This study was funded by the Novo Nordisk Foundation (#NNF17OC0027812), and by the National Institutes of Health (NIH) Common Fund SenNet program (U54AG075932).

## Introduction

Breast cancer is one of the most common forms of cancer worldwide and results in substantial mortality with 287,850 cases and 43,250 deaths annually in the United States alone.^1^ Numerous genetic and environmental factors contribute but the primary risk factor is age, with over 80% of cases occurring after the age of 50 and underlying the call to begin regular mammographic screening in all women by this age.^2^ Like many diseases of aging, breast cancer has been associated with cellular senescence^3–5^, where aged or damaged cells cease dividing but remain metabolically active.^6^ Senescence was first identified as a mechanism to limit proliferation of cells^7,8^ and later shown to operate with multiple pathways that respond to molecular damage to prevent excessive division, purportedly as a tumor suppressor mechanism.^9–11^ Paradoxically, senescent cells can also promote tumor development by emitting a variety of factors known as the senescence associated secretory phenotype (SASP), which produces an inflammatory state to signal immune clearance.^12–14^

Despite its relevance to numerous diseases including cancer, cellular senescence is poorly characterized in human tissues because of the lack of specific and universal biomarkers.^15^ While diverse markers are associated with senescence, they also report on other biological processes such as DNA damage, inflammation, and cell cycle, and there is no known marker to exclusively identify the senescent state. The best practices to overcome this challenge include staining for multiple markers simultaneously, although there is limited consensus regarding the right combination, which varies by tissue. This has led to recent initiatives such as SenNet by the NIH to map senescence in human tissues using the latest multiomics methods.^16^

Given the observation that senescent cells display altered nuclear morphology^17^, we recently showed that senescence can be detected using deep learning on tissue micrographs showing nuclear morphology.^18^ In the current study, we applied the nuclear senescence predictor (NUSP) to H&E-stained breast biopsy images from 4,411 healthy donors, of which 86 developed breast cancer at a later date, in order to investigate whether senescence is associated with future breast cancer development. Predicted senescence scores are generated for each nucleus and can be related to other nuclei by location and tissue type. Therefore, we analyzed the spatial distribution of predicted senescent cells by evaluating 32 million nuclei in breast tissue to explore how senescence arises in tissue and how it might relate to the risk of future breast cancer development.

## Methods

### Study design and population

Our retrospective cohort study utilized participants from the Komen Tissue Bank (KTB) at the Indiana University Simon Cancer Center, one of the largest biorepositories that collects, stores, and annotates breast tissue with a focus on female donors who exhibit no signs of breast disease.^19^ Tissue cores from the upper outer quadrant of either breast were acquired from consenting women using 10-gauge needles and immediately processed as snap-frozen tissues or using 10% formalin fixation and paraffin embedding (FFPE) or the PAXgene tissue preservation system prior to paraffin embedding (PFPE). Tissue was collected and processed by standardized KTB operating protocols and specimens were archived by the KTB.^20,21^ Donors completed questionnaires supplying demographic data and reproductive/medical histories at the time of donation as well as in follow-up questionnaires.

All KTB participants who underwent core biopsies for research purposes between 2009 and 2019 were eligible for this study. Digitized slide-based images of H&E stained non-malignant breast sections were obtained for all biopsied participants (n = 4,922), as were covariate data annotations including age at donation, race/ethnicity, parity, age at menarche, Body Mass Index (BMI), alcohol and smoking history, and family history of breast cancer. From these data, 5 year Gail scores were also calculated, representing the standard clinical predictor of breast cancer risk development and routinely used for all prevention studies. In participants with duplicate images (n = 326), we utilized the earlier biopsy image; those whose donation resulted from carrying a BRCA mutation (n = 15) and those with pre-existing breast cancer (n = 170) were excluded from the analysis, resulting in our study number of 4,411 participants. Participant characteristics according to case status are shown in Table 1. All women with breast cancer diagnosed at least 6 months after their last normal breast tissue biopsy donation were considered cases (n = 86).

**Table 1:**
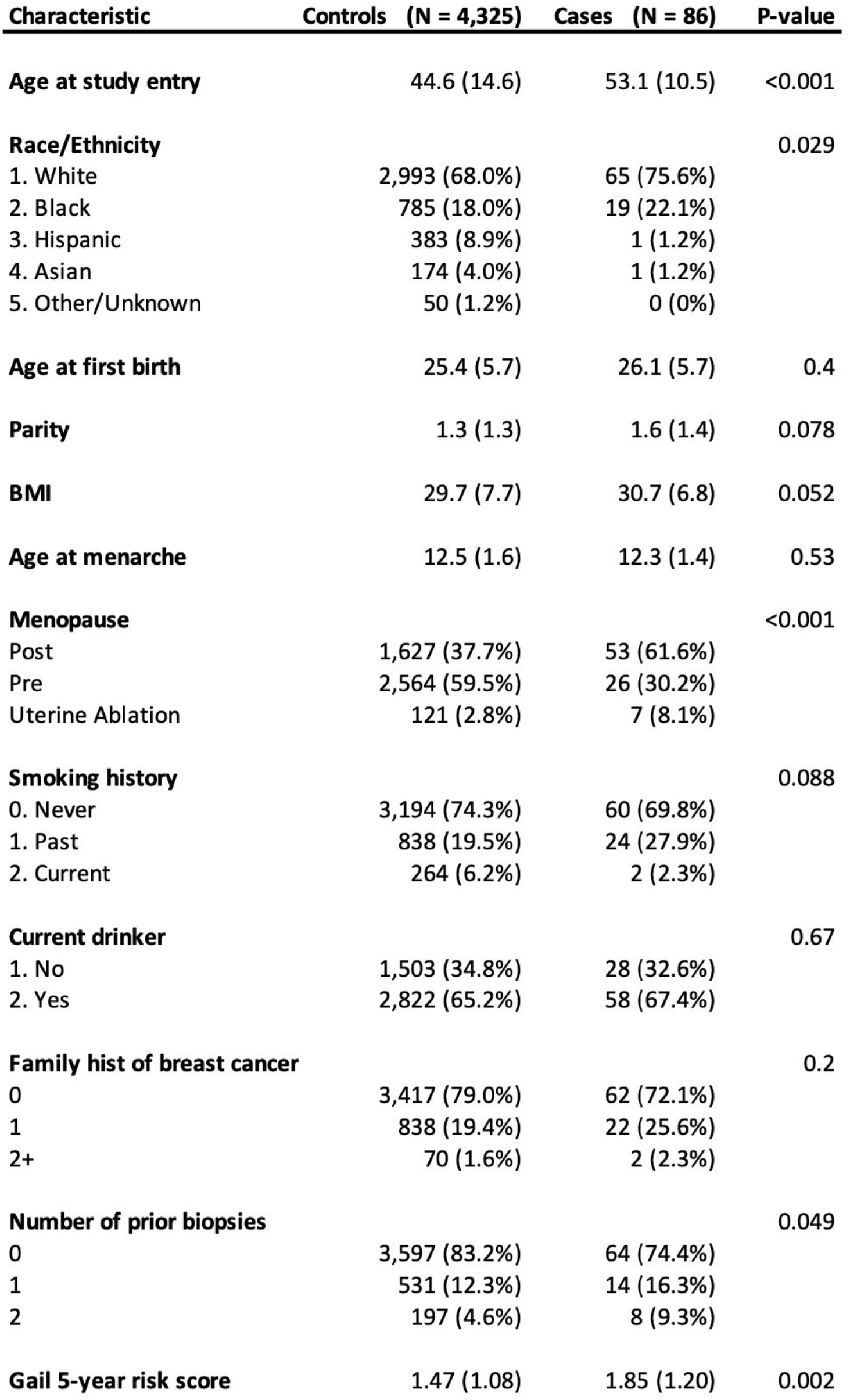
Outline of patient characteristics.

### Image Segmentation

H&E-stained whole slide images were provided by the Komen Tissue Bank as tiled tif (svs) files. To assist in the analysis, these large images were split into tiles of 1024 × 1024 pixels. Nuclei were identified using a U-Net model trained on 10 sample image tiles.^22^ Tissue was also segmented by training two U-Net models to detect epithelial and adipose regions, where each model was trained on 20 annotated tiles. Terminal duct lobular units (TDLUs) were segmented using a published model also based on U-Net.^23^ If a region was identified as TDLU, it was associated with that group only, and if not, it was then assessed to be non-TDLU epithelial or adipose tissue. If it did not match any of those groups, it was assumed to be stroma. A minority of nuclei were associated with both epithelial and adipose regions, and further examination indicated that these primarily represented non-adipose cells found adjacent to adipose cells, such as resident macrophages, endothelial cells, and others. This “both” group was excluded from the analysis due to its heterogeneity and the lower number of nuclei found in those regions.

### Senescence prediction

Cellular senescence was predicted from detected nuclei using the NUSP with models and methods developed by the authors.^15^ As described in the reference, all models were built on images from cell culture, which were size, background and intensity normalized to identify the nuclear outline and after training could predict senescence of normalized nuclei in tissue. The NUSP provides a standard unified (Uni) model for detecting senescence, but also offers several other models that were trained on senescence induced by multiple mechanisms, such as replicative exhaustion (RS), DNA damage (ionizing radiation, IR, and doxorubicin treatment, Doxo), mitochondrial dysfunction (antimycin-A treatment, Anti), and proteotoxicity (atazanavir-ritonavir treatment, ATVR). The Uni model was trained on all methods of induction together and has been shown to recognize all forms of senescence in vitro, whereas the individual models can offer superior performance in detecting senescence induced by specific methods. We applied all five individual models to determine which better captured the difference between cases and controls. Although some models captured significant effects in different tissues, several models showed similar results. We compared scores on a per-nucleus basis and observed a very high correlation between ATVR, Anti, and Doxo. While these three methods predict senescence induced by different mechanisms *in vitro*, our empirical results suggested that we can treat them together in breast tissue, leading to the training of a new model that integrates the three (AAD). This new AAD model was trained by combining the datasets used for the three individual models described in the publication (Anti, Atvr, and Doxo).^18^ Each senescence “model” is based on an ensemble of 10 models independently trained on the same data using the Xception architecture, and when applied, their outputs are combined to produce a mean score.

### Statistical analysis

Odds ratios (ORs) and 95% confidence intervals were calculated using logistic regression models to analyze the relationship between senescence scores and breast cancer incidence. Limitations in the regularity of follow-up donor data made it difficult to use time-to-event analyses. Breast tissue was segmented into fat, terminal duct lobular units (TDLUs), non-TDLU epithelium (epi), and stromal tissue for senescence scoring, and three senescence models were applied to the four tissue types. Results were examined for continuous (linear) association as well as by medians of senescent scores. Examination by quartiles was limited by the relatively small number of cases.

The two model results with the strongest senescent score/breast cancer risk associations were both found in fat tissue and were used in a cross-classification analysis looking at the risk for women with neither risk factor, having one of the risk factors, and having both risk factors. This analysis utilized logistic regression models to calculate ORs with 95% CI’s, comparing one or both risk factors to the reference group of having neither.

All models were adjusted for age at donation, race/ethnicity, parity, BMI, age at menarche, smoking history, alcohol intake, and family history of breast cancer. KTB changed their tissue fixation method during the interval being studied from formalin fixation and paraffin embedding (FFPE) to the PAXgene tissue preservation system prior to paraffin embedding (PFPE), resulting in 56.7% of slides using FFPE and 43.3% using PFPE. Because of potential batch issues created by the different techniques^24^, we performed batch adjustment by using the FFPE results as the target distribution and the PFPE sample results as the source distribution and used quantile normalization to minimize batch-specific differences.

Gail 5-year risk scores were generated for all study participants between the ages of 35 and 85 using source code from the “BCRA R package” found on the BCRAT website (https://dceg.cancer.gov/tools/risk-assessment/bcra). ORs using Gail 5-year risk scores by median were calculated by logistic regression to assess the relationship to breast cancer cases, and for use in cross-classification analyses to assess the risk for women in the higher median of 5-year Gail score who are also in the group with one of the two senescence score models demonstrating the highest risk association relative to participants with no risk factors as the reference group.

### Study approval

KTB participants and biopsy donors were initially recruited after providing written informed consent under a protocol approved by the Indiana University Institutional Review Board.

## Results

Following our hypothesis that cellular senescence may play a role in breast cancer development, we applied the NUSP to H&E-stained healthy breast tissue biopsy images from 4,411 KTB donors of various ages (Fig 1a). Notably, 86 individuals later went on to develop breast cancer (the average time to cancer diagnosis was 4.8 years). Although the NUSP has been shown to apply to multiple cell types and image preparation methods, it produces scores on different scales per tissue and imaging context. To reduce confounding factors and explore tissue-specific senescence, we segmented breast tissue into three primary tissue types (stromal, adipose, and epithelial). We also identified terminal duct lobular units, TDLUs, which are believed to be the primary site for origination of breast malignancies.^25^ By automatically recognizing tissue regions in each image, nuclei could be classified by the region where they were found. The NUSP was applied to generate senescence scores for each of the 32,857,482 nuclei recognized using multiple models, and their precise locations in the tissue images were tracked. Visualizing nuclei with high predicted senescence (over the 95th percentile) revealed spatial distribution throughout the tissues (Fig 1b). There was significant clustering in epithelial regions and TDLUs, although this may be due to their higher density of nuclei and limited spatial extent. We also noted frequent overlap of the prediction models (IR, RS, and AAD).

**Figure 1:**
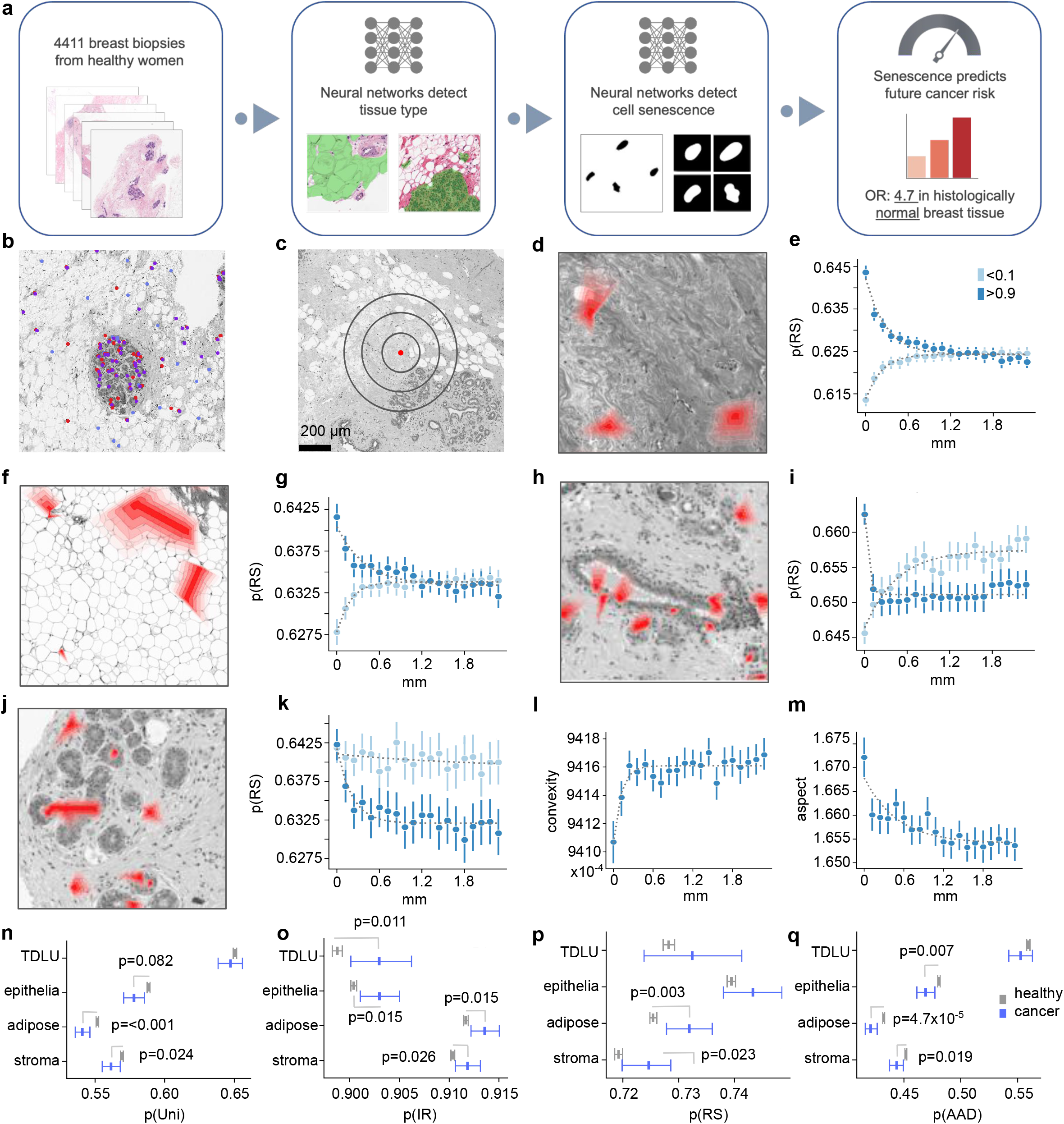
Cellular senescence organizes spatially and future cases show differences in tissues. **a**, Workflow for predicting senescence in tissue images. **b**, Representative histological images showing nuclei with high predicted senescence (>0.9). Models: ionizing radiation (IR) in red, replicative senescence (RS) in purple, and antimycin A, Atv/R and doxorubicin induced senescence (AAD) in blue. **c**, Representative image illustrating the distance cutoffs where senescence is measured. **d**, Representative image of stroma showing clusters (red) of RS-senescent nuclei. **e**, Mean RS scores of cells in stroma with increasing distance from senescent (score >0.9) and non-senescent (score <0.1) cells (exponential curve fit, >0.9: R^2^=0.95, <0.1: R^2^=0.98). **f**, Adipose tissue sample, showing regions of high RS-scoring senescent nuclei in red. **g**, Mean RS scores of cells in adipose tissue with increasing distance from senescent (score >0.9) and non-senescent (score <0.1) cells (exponential curve fit, >0.9: R^2^=0.87, <0.1: R^2^=0.97). **h**, Epithelia sample, showing regions of high RS-scoring senescent nuclei in red. **i**, Mean RS scores of epithelial cells with increasing distance from senescent (score >0.9) and non-senescent (score <0.1) cells (exponential curve fit, >0.9: R^2^=0.89, <0.1: R^2^=0.90). **j**, Terminal ductile lobular unit (TDLU) sample, showing regions of high RS-scoring senescent nuclei in red. **k**, Mean RS scores of cells in TDLUs with increasing distance from senescent (score >0.9) and non-senescent (score <0.1) cells (exponential curve fit, >0.9: R^2^=0.86, <0.1: R^2^=0.14). **l**, Mean convexity of nuclei in adipose tissue near high >0.9 percentile IR-scoring nuclei (exponential curve fit, R^2^=0.83). **m**, Mean aspect of nuclei in adipose tissue near high >0.9 percentile IR-scoring nuclei (exponential curve fit, R^2^=0.76). Mean senescence scores for individuals per tissue for cases vs. controls, using the Uni (**n**), IR (**o**), RS (**p**) and AAD (**q**) models (mean ± 95% CI).

Previous work has suggested that senescent cells can induce senescence in neighboring cells.^26^ We evaluated the spatial distribution of senescence by classifying nuclei by percentile and examining mean senescence score (normalized per individual) for nearby nuclei by distance (Fig. 1c). For all three models and the three main tissue types, nuclei were surrounded by other nuclei with similar scores, indicating clustering where senescent nuclei were found near other high-senescence-scoring nuclei. As the distance from senescent nuclei increased, the mean senescence score decreased, indicating an increasing rate of non-senescent nuclei (Fig. 1c-k, Ext Fig. 1a-c, e-h). Likewise, with a greater distance from non-senescent nuclei, the mean senescence increases, indicating that more senescent nuclei are found (Fig. 1c-i, Ext Fig. 1a-h). While this pattern is largely consistent among models and tissues, we found that the IR model suggests that senescent TDLU nuclei are more likely to be surrounded by lower-senescence-scoring nuclei (Ext Fig 1d). In addition, the AAD model suggests that senescent TDLU nuclei are found alongside senescent nuclei, but there is reduced senescence outside its immediate neighborhood (Ext Fig 1h). For other tissue types (stroma, fat, and epithelium), the clustering of high senescence scoring and low senescence scoring nuclei suggests causal factors induce senescence in groups of cells or that senescent cells propagate their state through contact or by SASP inflammatory factors, forming regions of increased senescence. Observing the steep decline in mean senescence with distance, we fit an inverse exponential curve, obtaining good fits with high R^2^ (Fig. 1e,g, Ext Fig. 1a,c,e,g,h). The spatial distribution of senescence near high-scoring senescent nuclei resembles exponential decay, further supporting the notion that one or more paracrine SASP factors propagate senescence by local diffusion.

Considering that cell density could influence senescence and vice versa, we tested if the mean count of nuclei differs in the vicinity of high and low scoring nuclei. The AAD model suggested higher density of cells in the neighborhood of high-senescence-scoring nuclei (Ext Fig 1i). The RS model did not show differences, and the IR model showed higher counts for epithelia but lower counts for adipose cells (Ext Fig 1j,k). Although speculative, these results could indicate that the higher density for senescent nuclei is related to an inflammatory response, where immune cells are recruited by factors in the SASP.

To better characterize predicted senescence, we investigated morphological metrics near high-senescence scoring nuclei. We evaluated convexity and aspect ratio, which were shown to indicate senescence (Fig 1l,m).^18^ Indeed, we found substantially higher aspect and reduced convexity, both indicating senescence, in the closest cells to high senescent scoring cells. These patterns are consistent for all tissue types and both the IR and RS models, indicating clustering of senescent cells.

To investigate if senescence relates to cancer formation, we looked for differences in the 86 cases that developed cancer within our cohort. Notably, future cancer cases appeared to have a smaller number of nuclei in all their normal tissue types, with a small but significant difference in adipose, epithelia, and stroma, and a larger difference in TDLU (Ext Fig. 1l). This could be related to the senescence response, with reduced immune recruitment, a reduced protective effect of senescence, or other subtle changes to tissue.

Since we observed spatial differences in predicted senescence for multiple tissue types, we investigated whether tissue specific senescence could be a prognostic marker for cancer. Using the unified model (Uni), we found that the cases appeared significantly different from the controls, with cases having a lower mean predicted senescence in all three tissue types (Fig 1n). Senescence appears to have a protective effect in multiple tissues, reducing the risk of developing cancer. Importantly, biopsies were taken 57.5 months before diagnosis, on average, suggesting that the predicted rate of senescence may be a critical indicator of the risk for developing malignancy.

We also applied the senescence prediction models trained on several specific types of stress to evaluate whether different types of senescence can influence carcinogenesis. Strikingly, we observed significant differences in mean senescence for all cell types with IR, for adipose and stroma with RS, and for epithelial, adipose, and stromal tissue with AAD (Figs. 1o-q). While the AAD model shows a protective role for senescence where cancer cases tend to have lower scores, the IR and RS models suggest that higher senescence scores increase the risk of cancer, indicating that different types of senescence may impact carcinogenesis.

After evaluating overall senescence per tissue per individual and adjusting for batch effects, we found a statistically significant decrease in the risk of developing breast cancer for individuals with high AAD scores in fat (above the median) compared to individuals with low scores (below the median), OR=0.57 ([0.36-0.88], p=0.013), indicating that women with the lower half of senescence scores had a substantially higher risk of cancer development than those in the upper half (Fig 2a, Ext. Table 1 for unadjusted results). Conversely, with the IR model we found significantly increased risk of cancer development for individuals with higher predicted senescence in fat tissue (OR=1.71, [1.10-2.68], p=0.019). This was also found for the IR model in stromal tissue (OR=1.59 [1.01-2.49]). Although other model and tissue results did not reach statistical significance, results of the IR and RS models on all 4 tissue types were above 1, while 3 of 4 tissue types for the AAD model were below 1. These results suggest that some forms of senescence can have a protective effect that can substantially reduce cancer risks, while other forms of senescence might promote cancer development.

**Figure 2:**
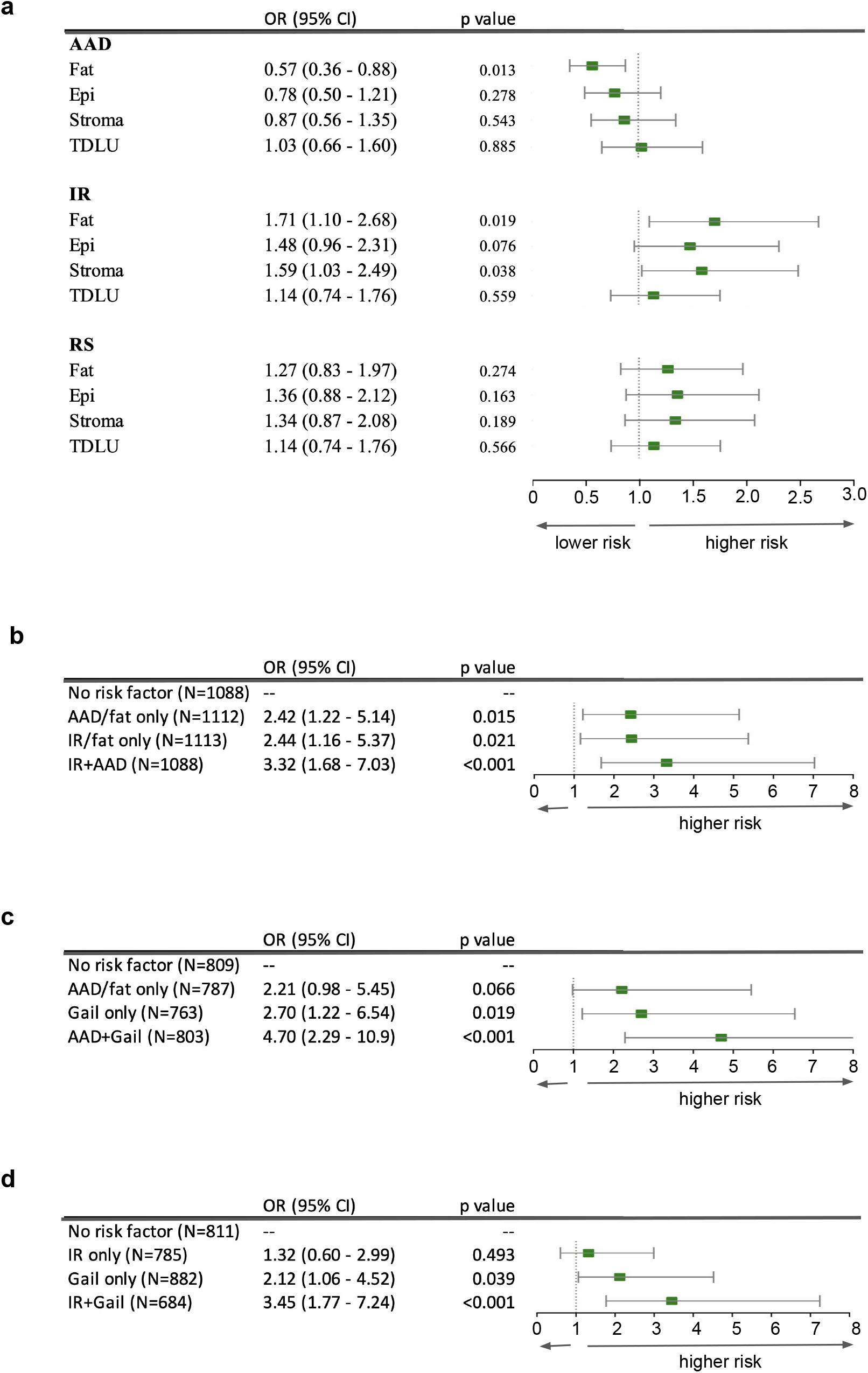
Senescence indicates risk of developing cancer. **a**, Estimated risk by model and tissue type with p-value and 95% confidence interval. **b**, Cross-classification analysis showing OR by combining the IR and AAD models for fat. **c**, Cross-classification analysis showing OR by combining AAD for fat with 5 year Gail scores. **d**, Cross-classification analysis showing OR by combining IR for fat with 5 year Gail scores.

Cross-classification analyses suggest that the risk indicated by each model was largely independent (Fig 2b). Individuals with both low AAD/fat scores and high IR/fat scores had OR=3.32 ([1.68-7.03], p<0.001), suggesting that multiple forms of senescence relate to risk differently, with the AAD model capturing a protective effect and the IR model being associated with increased risk. We also generated 5-year Gail scores for all individuals and performed a similar analysis. Remarkably, individuals with both high Gail scores and low AAD/fat scores had an OR of 4.70 ([2.29-10.90], p<0.001; Fig 2c) while both high Gail scores and high IR/fat scores yielded an OR=3.45 ([1.77-7.24], p<0.001; Fig 2d). Importantly, these results indicate little overlap in the risk derived from the different predicted senescence models and the Gail score.

## Discussion

In this study, we applied deep learning models to predict senescence per nucleus in H&E-stained micrographs of healthy breast tissues from 4,411 individuals. Remarkably, the NUSP identified significant differences in the mean senescence score for the 86 individuals in our cohort who were later diagnosed with breast cancer at an average interval of 4.8 years after their non-malignant breast biopsies. Our risk analysis suggested that individuals in the higher median of predicted senescence scores for adipose tissue using the AAD model had a reduced risk of developing cancer (OR = 0.57, [0.36-0.88], p=0.013), while individuals in the higher median of the IR model had an elevated risk (OR = 1.71, [1.10-2.68], p=0.019). Individuals with both risk factors have OR=3.32 ([1.68-7.03], p<0.001), which exceeds the clinical gold standard in the field, the Gail model, giving OR=2.33 for the upper median of scores. Combining a single senescence risk factor (AAD/fat) with the Gail model resulted in an OR=4.70 ([2.29-10.90], p<0.001), considerably higher than that predicted by the Gail score alone. This could likely be further improved by utilizing multiple senescence models with Gail scores to enhance the prediction of breast cancer risk. In summary, our analysis suggests dual roles for senescence in maintaining tissue health while also being associated with malignancy, and we introduce new senescence-based risk models that can be applied directly to H&E-stained images.

Individual models trained on various senescence inducers indicated that senescence can simultaneously promote and protect against cancer, depending on the form of senescence. The IR model, trained on senescent cells with severe DNA damage induced by radiation, showed higher senescence in the stroma and fat for those who developed cancer, whereas the AAD model showed a protective effect from senescence in those same tissue constituents. The positive relationship between the IR model and cancer could be due to the effect of the pro-inflammatory SASP, which has been shown to be particularly prominent in IR-induced cells.^29^ Senescence arising from significant DNA damage and captured by the IR model could also be related to a higher rate of mutations that could contribute to malignant transformation. Conversely, the AAD model was trained on senescence induced by diverse drug treatments, with antimycin inducing mitochondrial dysfunction and ATV/r inducing proteotoxic stress. Increased rates of these types of senescence could provide a protective effect by controlling proliferation without mutational load and SASP effects. The dual roles of senescence in promoting and controlling cancer are widely recognized^30,31^, and our study provides evidence for both roles using high-precision machine learning methods to analyze a large cohort. We speculate that the IR model is capturing a stronger senescence phenotype with a more developed SASP, while the AAD model, having been trained on diverse drug treatments, may reflect a weaker or earlier senescence phenotype with a reduced SASP^32^.

The use of 4000+ women from the KTB with biopsy images and associated covariates was a major strength of the study, as all KTB women underwent biopsies at the time of enrollment, and the development of breast cancer occurred after biopsy in all study participants. The relatively small number of cases (n = 86) was a limitation, and although the effect size was large enough to find statistical significance in breast cancer cases as a whole, it did prevent subcategorization by breast cancer type or hormone receptor status. There is also the possibility of selection bias, as not all women are willing to undergo breast biopsy on a voluntary basis, and the race/ethnicity of the KTB participants may limit the generalizability to the overall population or to under-represented subgroups.

Our spatial analysis also uncovered several notable aspects of cellular senescence in breast tissues. The SASP induces changes through paracrine factors, and for all three major tissue types, senescent nuclei were on average found near similar nuclei, and the rate of senescent nuclei declined with distance, suggesting clusters of senescence. We also showed that senescent epithelial (including TDLU) nuclei exist in regions of higher density, which may suggest that higher density induces senescence or that these higher-density regions are more susceptible to senescence. Comparing the density for cases to controls revealed a lower density for cases across all three tissue types along with TDLUs, supporting the notion that at least locally senescence serves as a barrier to cancer.

In conclusion, the rate of senescence predicted from nuclear morphology in H&E-stained healthy breast tissue images is a strong risk-predictor of breast cancer development. This relationship is found for individuals overall and per tissue. In our cohort, the combination of multiple models greatly improved risk-prediction compared with the current clinical benchmark, the Gail score. Notably, our deep learning senescence models predict breast cancer risk orthogonally to the Gail model and achieve nearly twice the OR when combined with the Gail model. In sum, high-precision predicted senescence scores provide new insights into breast cancer risk and may be applied in the clinical assessment and screening of women with non-malignant breast biopsies that are not otherwise useful and may be potentially misleading with respect to future breast cancer risk assessment.

## Data Availability

All data produced in the present study are available upon reasonable request to the authors

## Author Contributions

IH wrote the manuscript, applied the deep learning models, and analyzed the data. MP conceived, designed, and executed the study, supervised the analysis, and edited the manuscript; SF analyzed the KTB data; JH, SR, and JC provided access to necessary KTB images and covariates; AT managed clinical samples and edited the manuscript; EV advised the project; CB contributed to the KTB collaboration, consulted on the study results, and edited the manuscript; MSK supervised the project and edited the manuscript.

## Acknowledgments

This research was supported by the Novo Nordisk Foundation Challenge Programme (NNF17OC0027812), the Nordea Foundation (02-2017-1749), the Neye Foundation, the Lundbeck Foundation (R324-2019-1492), the Ministry of Higher Education and Science (0238-00003B), VitaDAO, Insilico Medicine and the U54 AG075932 grant from the National Institutes of Health. The funders had no role in the study design, data collection and analysis, decision to publish, or manuscript preparation.

The authors would like to acknowledge Renata Cora, who provided annotation of TDLU’s that was used to train segmentation detectors.

Data from the Susan G. Komen Tissue Bank at the IU Simon Cancer Center were used in this study. We thank contributors, including Indiana University who collected data used in this study, as well as donors and their families, whose help and participation made this work possible.

## Legend

**Ext Figure 1:**
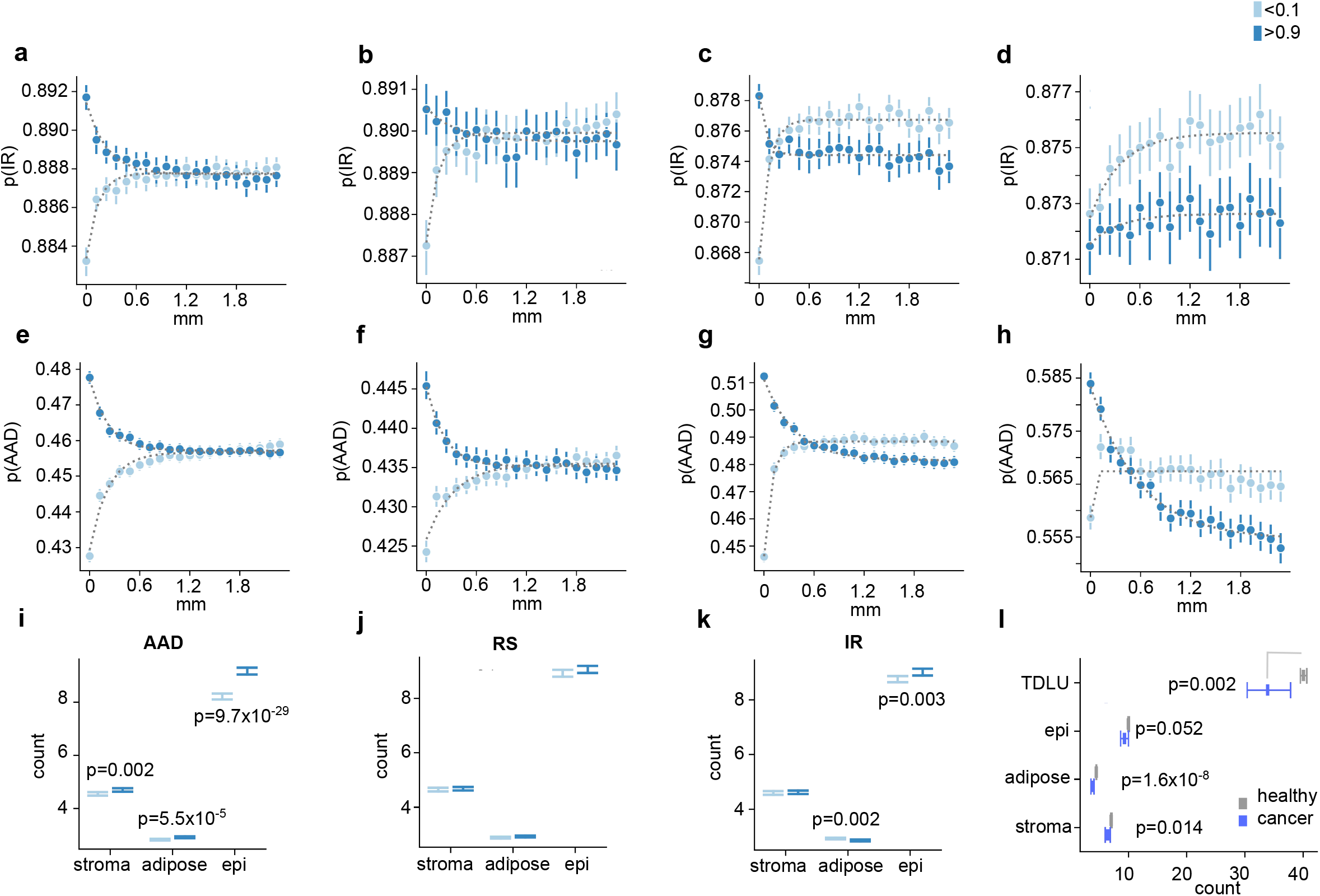
Additional spatial analysis and nuclei counts by tissue. **a**, Mean IR scores of cells in stroma with increasing distance from senescent (score >0.9) and non-senescent (score <0.1) cells (exponential curve fit, >0.9: R^2^=0.93, <0.1: R^2^=0.94). **b**, Mean IR scores of cells in adipose with increasing distance from senescent (score >0.9) and non-senescent (score <0.1) cells (exponential curve fit, >0.9: R^2^=0.54, <0.1: R^2^=0.88). **c**, Mean IR scores of epithelial cells with increasing distance from senescent (score >0.9) and non-senescent (score <0.1) cells (exponential curve fit, >0.9: R^2^=0.77, <0.1: R^2^=0.96). **d**, Mean IR scores of cells in TDLUs with increasing distance from senescent (score >0.9) and non-senescent (score <0.1) cells (exponential curve fit, >0.9: R^2^=0.39, <0.1: R^2^=0.82). **e**, Mean AAD scores of cells in stroma with increasing distance from senescent (score >0.9) and non-senescent (score <0.1) cells (exponential curve fit, >0.9: R^2^=0.99, <0.1: R^2^=0.96). **f**, Mean AAD scores of cells in adipose with increasing distance from senescent (score >0.9) and non-senescent (score <0.1) cells (exponential curve fit, >0.9: R^2^=0.97, <0.1: R^2^=0.90). **g**, Mean AAD scores of epithelial cells with increasing distance from senescent (score >0.9) and non-senescent (score <0.1) cells (exponential curve fit, >0.9: R^2^=0.98, <0.1: R^2^=0.90). **h**, Mean AAD scores of cells in TDLUs with increasing distance from senescent (score >0.9) and non-senescent (score <0.1) cells (exponential curve fit, >0.9: R^2^=0.98, <0.1: R^2^=0.38). **I**, Count of nuclei near high >0.9 and low <0.1 scoring nuclei using the AAD model. **j**, Count of nuclei near high >0.9 and low <0.1 scoring nuclei using the RS model. **k**, Count of nuclei near high >0.9 and low <0.1 scoring nuclei using the IR model. **l**, Mean count of nuclei per 45238 μm^2^ for each tissue type by cases and controls (mean ± 95% confidence interval (CI)).

---

**Ext Table 1:** Unadjusted risk results.

| Characteristic | Stroma |  |  | Epi |  |  | TDLU |  |  | Fat |  |  |
| --- | --- | --- | --- | --- | --- | --- | --- | --- | --- | --- | --- | --- |
|  | OR | 95% CI | p-value | OR | 95% CI | p-value | OR | 95% CI | p-value | OR | 95% CI | p-value |
| Continuous | 0.92 | 0.85,1.00 | 0.044 | 0.93 | 0.88,0.99 | 0.018 | 0.98 | 0.94,1.02 | 0.32 | 0.86 | 0.78,0.93 | <0.001 |
| 1. 1st Quartile | — | — |  | — | — |  | — | — |  | — | — |  |
| 2. 2nd Quartile | 0.92 | 0.52,1.61 | 0.75 | 0.78 | 0.44,1.37 | 0.39 | 0.49 | 0.25,0.92 | 0.031 | 0.50 | 0.28,0.89 | 0.021 |
| 3. 3rd Quartile | 0.40 | 0.19,0.80 | 0.012 | 0.39 | 0.19,0.77 | 0.009 | 0.71 | 0.38,1.29 | 0.27 | 0.44 | 0.23,0.80 | 0.008 |
| 4. 4th Quartile | 0.57 | 0.30,1.06 | 0.080 | 0.57 | 0.30,1.06 | 0.081 | 0.74 | 0.40,1.35 | 0.33 | 0.27 | 0.13,0.54 | <0.001 |
| Quartiles |  |  |  |  |  |  |  |  |  |  |  |  |
| 1. 1<sup>st</sup> Quartile | — | — |  | — | — |  | — | — |  | — | — |  |
| 2. 2<sup>nd</sup>-4<sup>th</sup> Quartiles | 0.63 | 0.39,1.02 | 0.054 | 0.58 | 0.37,0.94 | 0.025 | 0.64 | 0.40,1.04 | 0.062 | 0.40 | 0.26,0.65 | <0.001 |

RS Model
| Characteristic | Stroma |  |  | Epi |  |  | TDLU |  |  | Fat |  |  |
| --- | --- | --- | --- | --- | --- | --- | --- | --- | --- | --- | --- | --- |
|  | OR | 95% CI | p-value | OR | 95% CI | p-value | OR | 95% CI | p-value | OR | 95% CI | p-value |
| Continuous | 1.09 | 0.99,1.20 | 0.080 | 1.05 | 0.96,1.14 | 0.28 | 1.03 | 0.97,1.10 | 0.28 | 1.16 | 1.04,1.29 | 0.009 |
| 1. 1st Quartile | — | — |  | — | — |  | — | — |  | — | — |  |
| 2. 2nd Quartile | 1.31 | 0.64,2.72 | 0.46 | 0.79 | 0.38,1.62 | 0.52 | 0.97 | 0.51,1.84 | 0.92 | 1.16 | 0.56,2.44 | 0.68 |
| 3. 3rd Quartile | 1.99 | 1.04,3.96 | 0.043 | 1.56 | 0.85,2.96 | 0.15 | 0.91 | 0.47,1.74 | 0.76 | 1.61 | 0.82,3.27 | 0.17 |
| 4. 4th Quartile | 1.71 | 0.87,3.45 | 0.12 | 1.32 | 0.69,2.56 | 0.40 | 1.17 | 0.64,2.17 | 0.61 | 2.14 | 1.13,4.26 | 0.023 |
| Quartiles |  |  |  |  |  |  |  |  |  |  |  |  |
| 1. 1<sup>st</sup> Quartile | — | — |  | — | — |  | — | — |  | — | — |  |
| 2. 2<sup>nd</sup>-4<sup>th</sup> Quartile | 1.66 | 0.95,3.11 | 0.089 | 1.21 | 0.72,2.16 | 0.48 | 1.02 | 0.62,1.73 | 0.95 | 1.62 | 0.93,3.03 | 0.11 |

IR Model
| Characteristic | Stroma |  |  | Epi |  |  | TDLU |  |  | Fat |  |  |
| --- | --- | --- | --- | --- | --- | --- | --- | --- | --- | --- | --- | --- |
|  | OR | 95% CI | p-value | OR | 95% CI | p-value | OR | 95% CI | p-value | OR | 95% CI | p-value |
| Continuous | 1.70 | 1.18,2.49 | 0.005 | 1.31 | 1.04,1.64 | 0.019 | 1.24 | 1.05,1.45 | 0.009 | 1.63 | 1.18,2.27 | 0.004 |
| 1. 1st Quartile | — | — |  | — | — |  | — | — |  | — | — |  |
| 2. 2nd Quartile | 1.45 | 0.71,3.02 | 0.31 | 1.16 | 0.58,2.33 | 0.68 | 1.25 | 0.63,2.52 | 0.52 | 2.26 | 1.11,4.81 | 0.027 |
| 3. 3rd Quartile | 2.09 | 1.06,4.25 | 0.036 | 1.42 | 0.71,2.87 | 0.31 | 1.36 | 0.69,2.73 | 0.32 | 2.01 | 0.96,4.35 | 0.067 |
| 4. 4th Quartile | 2.49 | 1.29,5.02 | 0.008 | 2.08 | 1.12,4.01 | 0.023 | 1.64 | 0.86,3.22 | 0.14 | 3.39 | 1.71,7.10 | <0.001 |
| Quartiles |  |  |  |  |  |  |  |  |  |  |  |  |
| 1. 1<sup>st</sup> Quartile | — | — |  | — | — |  | — | — |  | — | — |  |
| 2. 2<sup>nd</sup>-4<sup>th</sup> Quartile | 1.97 | 1.12,3.70 | 0.025 | 1.53 | 0.90,2.77 | 0.14 | 1.41 | 0.82,2.59 | 0.23 | 2.50 | 1.38,4.93 | 0.004 |

